# Application and optimization of RT-PCR in diagnosis of SARS-CoV-2 infection

**DOI:** 10.1101/2020.02.25.20027755

**Authors:** Xiaoshuai Ren, Yan Liu, Hongtao Chen, Wei Liu, Zhaowang Guo, Yaqin Zhang, Chaoqun Chen, Jianhui Zhou, Qiang Xiao, Guanmin Jiang, Hong Shan

**Affiliations:** Clinical Laboratory, The Fifth Affiliated Hospital, Sun Yat-Sen University. Zhuhai, 519000, China; Department of Radiology, The Fifth Affiliated Hospital of Sun Yat-sen University, No. 52 Meihua Dong Road, Zhuhai 519000, Guangdong Province, People’s Republic of China; Central Laboratory, The Fifth Affiliated Hospital of Sun Yat-sen University, Zhuhai, Guangdong, China; Key Laboratory of Biomedical Imaging of Guangdong Province, Guangdong Provincial Engineering Research Center of Molecular Imaging, The Fifth Affiliated Hospital of Sun Yat-sen University, Zhuhai, 519000, Guangdong, China

**Author notes:** **Corresponding author:** Hong Shan, PhD. Guanmin Jiang, PhD **Address for correspondence:** Hong Shan, Ph.D., Professor, Mailing address: Key Laboratory of Biomedical Imaging of Guangdong Province, Guangdong Provincial Engineering Research Center of Molecular Imaging, The fifth Affiliated Hospital of Sun Yat-sen University, No. 52 Meihua Dong Road, Zhuhai 519000, Guangdong Province, People’s Republic of China., Guanmin Jiang, Ph.D., Professor, Mailing address: Clinical Laboratory, The fifth Affiliated Hospital, Sun Yat-Sen University. Zhuhai, 519000, China. These authors contributed equally to this article (Xiaoshuai Ren, Yan Liu, Hongtao Chen). These senior authors contributed equally to this article.

**Keywords:** COVID-19, SARS-CoV-2, RT-PCR, chest CT

## Abstract

**Background:** Coronavirus Disease 2019 (COVID-19) caused by Severe acute respiratory syndrome coronavirus 2 (SARS-CoV-2) has become a global threat to public health. Aiming to construct an efficient screening pattern, we comprehensively evaluated the performances of RT-PCR and chest CT in diagnosing COVID-19.

**Methods:** The records including demographics, RT-PCR, and CT from 87 confirmed COVID-19 cases and 481 exclusion cases were collected. The diagnostic accuracy of the pharyngeal swab RT-PCR, CT, combination with the second pharyngeal swab RT-PCR or with CT were evaluated individually. Besides, all the stool RT-PCR results were plotted by time to explore the value of stool RT-PCR.

**Findings:** Combination of RT-PCR and CT has the higher sensitivity (91.9%,79/86) than RT-PCR alone (78.2%,68/87) or CT alone (66.7%, 54 of 81) or combination of two RT-PCR tests (86.2%,75/87). There was good agreement between RT-PCR and CT (kappa-value, 0.430). In 34 COVID-19 cases with inconsistent results, 94.1% (n=32) are mild infection, 62.5% of which (20/32) showed positive RT-PCR. 46.7% (35/75) COVID-19 patients had at least one positive stool during the course. Two cases had positive stool earlier than the pharyngeal swabs. Importantly, one patient had consecutive positive stool but negative pharyngeal swabs.

**Interpretation:** Combination of RT-PCR and CT with the highest sensitivity is an optimal pattern to screen COVID-19. RT-PCR is superior to CT in diagnosing mild infections. Stool RT-PCR should be considered as an item for improving discovery rate and hospital discharge. This study shed light for optimizing scheme of screening and monitoring of SARS-CoV-2 infection.

**Funding:** This work was supported by the National Natural Science Foundation of China (No. 81502104), National Program on Key Basic Research Project (No. 2018YFC0910600),the Nature Science Foundation of Guangdong Province, China (Grant No: 2017A030313771 and 2020A151501001) and the Young Teachers Nurturing Program of Sun Yat-Sen University (Grant No:17ykpy62)

## Introduction

In early December 2019, the first pneumonia cases of unknown origins were identified in Wuhan city, Hubei province, China^1^. On Jan 7, a novel coronavirus was discovered using high-throughput sequencing in the throat swab sample of a patient, and is currently named SARS-CoV-2(previously known as 2019-nCoV)on February 11, 2020 by ICTV^2,3^. The initial defined cases of COVID-19, were epidemiologically linked to the human seafood market in Wuhan, Although later more and more COVID-19 were found without exposure the market but with a history to Wuhan or contact with the patient of COVID-19 pneumonia confirmed^2,4,5^. Current epidemiologic data indicate the person-to-person transmission of SARS-CoV-2 in hospital and family settings^2,6,7^. As of February 17, 2020, more than 71,000 laboratory-confirmed and 1,770 death cases have been documented in China and in other countries worldwide (including the USA, German, japan and South Korea)^8,9^. The mortality rate of SARS-CoV-2 was around 2%. The WHO has recently declared the SARS-CoV-2 a public health emergency of international concern^10^. Thus, diagnostic tests specific for this infection are urgently needed for confirming suspected cases, screening patients and conducting virus surveillance.

Identification of pathogens mainly includes virus isolation and viral nucleic acid detection. According to the traditional Koch’s postulates, virus isolation is the gold standard for virus diagnosis in the laboratory. Thus, based on SARS-CoV-2 possesses a strong capability to infect humans, CDC recommends that clinical virology laboratories should not attempt viral isolation from specimens collected from COVID-19 patients under investigation. Because SARS-CoV-2 is a newly discovered virus, the spectrum of the available diagnostic tools is tight. In the early stage, SARS-CoV-2 has been detected in human clinical specimens by next-generation sequencing, cell culture, and electron microscopy^11^. Further development of accurate and rapid methods to identify this emergency respiratory pathogen is still needed.

Then the full genome sequence of SARS-CoV-2 (29870-bp, excluding the poly (A) tail) in GenBank (accession number MN908947) was released quickly on January 10, 2020, which is more than 82% identical to those of SARS-CoV and bat SARS-like coronavirus (SL-CoV)^12^. On the basis of analysis of this complete genomes obtained in this study, several laboratories developed molecular detection tools based on targeting ORF1ab, RNA-dependent RNA polymerase (RdRp) gene N, and E regions of viral spike genes^13-15^. And then the rapid identification of this novel coronavirus is attributed to recent advances in the detection of SARS-CoV-2, including RT-PCR, real-time reverse transcription PCR (rRT-PCR), reverse transcription loop-mediated isothermal amplification (RT-LAMP), and microarray-based assays. At present, RT-PCR is a widely used detection technique for SARS-CoV-2 and several marketed nucleic acid detection kits for using in clinic^14^. Currently, the standard of reference for the COVID-19 pneumonia diagnosis is a positive result in nucleic acid detection assay for the upper and lower respiratory tract specimens and blood, respiratory tract specimens were including nasal and pharyngeal swab specimens, sputum, and bronchoalveolar lavage fluid. And the patients confirmed with the COVID-19 pneumonia had 2 or 3 continuous negative RT-PCR results for nasopharyngeal and throat swab specimens can discharge from hospital. However, the scholar around china indicated that cases of COVID-19 that had 2 or 3 continuous negative RT-PCR results for nasopharyngeal and throat swab specimens before finally laboratory-confirmed[13]. And currently, several reports has reported the positive RT-PCR results for stool of COVID-19 patients^16,17^. Based on the infected patients can potentially shed the SARS-CoV2 through respiratory and fecal-oral routes, The value of RT-PCR results for stool in early diagnosis and monitor of SARS-CoV-2 infection will be study.

Fever, cough and dyspnea were the most common symptoms in patients with COVID-19 pneumonia. A manifestation similar of those of two other disease caused by coronaviruses, severe acute respiratory syndrome (SARS) and Middle East respiratory syndrome (MERS)^18-20^. CT is an important method in the diagnosis of lung lesions, and the radiological changes in the lungs of COVID-19 patients has been characterized^21^. Zhong et al. reported that of 840 COVID-19 patients who underwent CT on admission, around 76.4% manifested abnormal CT imaging features and usually exhibited typical radiological finding of the ground-glass opacity (50%) or bilateral patchy shadowing (46%)^22^. Based on the “Diagnosis and Treatment Guideline for New Coronavirus Pneumonia (the fifth edition), China”, CT scan were used as the clinical diagnostic criteria for COVID-19 pneumonia, but strictly limited in Hubei Province^23^. However, the specificity of chest CT is relatively low,alone could not distinguish the SARS-CoV-2 infection from other pathogens well.

SARS-CoV-2 causes extensively outbreak in cold winter. In this season, many other pathogens causing pneumonia also become prevalent, even including many viral agent. The infectious diseases share some common characteristics in signs, symptoms and laboratory findings. Therefor it is difficult to differentiate COVID-19 suffers from other pneumonia patients purely depending to the manifestation or routine examination. Therefore, an precision screening scheme is urgent to be employed. High sensitive test is pivotal to avoiding secondary transmission by missed diagnosed cases. Meanwhile, the positive predictive value also should be counted, for a number of false positive would bring out not only occupation and cost of healthcare resource, but also increasing infection risk of suspected cases isolated in hospital. In this study, we performed a retrospective study in the 568 cases and compare the efficacy of RT-PCR and CT diagnostic approaches in COVID-19 diagnosis, and to provide evidence for future strategic diagnosis in regions outside Hubei Province.

## Methods

### Data sources

For this retrospective, single center study, we recruited 584 patients from Jan 17 to Feb 11, 2020, at The Fifth Affiliated Hospital of Sun Yat-sen University in Zhuhai, China, which is a designated infectious hospital. During this period, RT-PCR and chest CT was performed for consecutive patients including the local residents of Wuhan, outside of Wuhan did have a recent travel to Wuhan or contact with people with fever or respiratory symptoms from Wuhan, or had fever or acute respiratory symptoms of unknown cause. Of the 16 patients recruited as of Feb 11, had a suspected diagnosis and were therefore excluded in this study. 87 patients, who were diagnosed as having COVID-19 and 481 patients exclusion COVID-19 according to WHO interim guidance, were enrolled in this study. The performances of the first RT-PCR detection in pharyngeal swabs and chest CT were evaluated by sensitivity, specificity, youden’s index et al. Then the performances of combination of the second RT-PCR, or chest CT were also calculated. Agreement between the two method was analyzed using McNemar Chi-squared test. Finally the all RT-PCR results from pharyngeal and stool were plotted by time to explore the value of stool nucleic detection (**Fig 1**). The severity of COVID-19 pneumonia was defined based on the international guidelines for community-acquired pneumonia^24^. Laboratory and CT characteristics data were obtained with standard data collection forms from electronic medical records.

**Figure 1.**
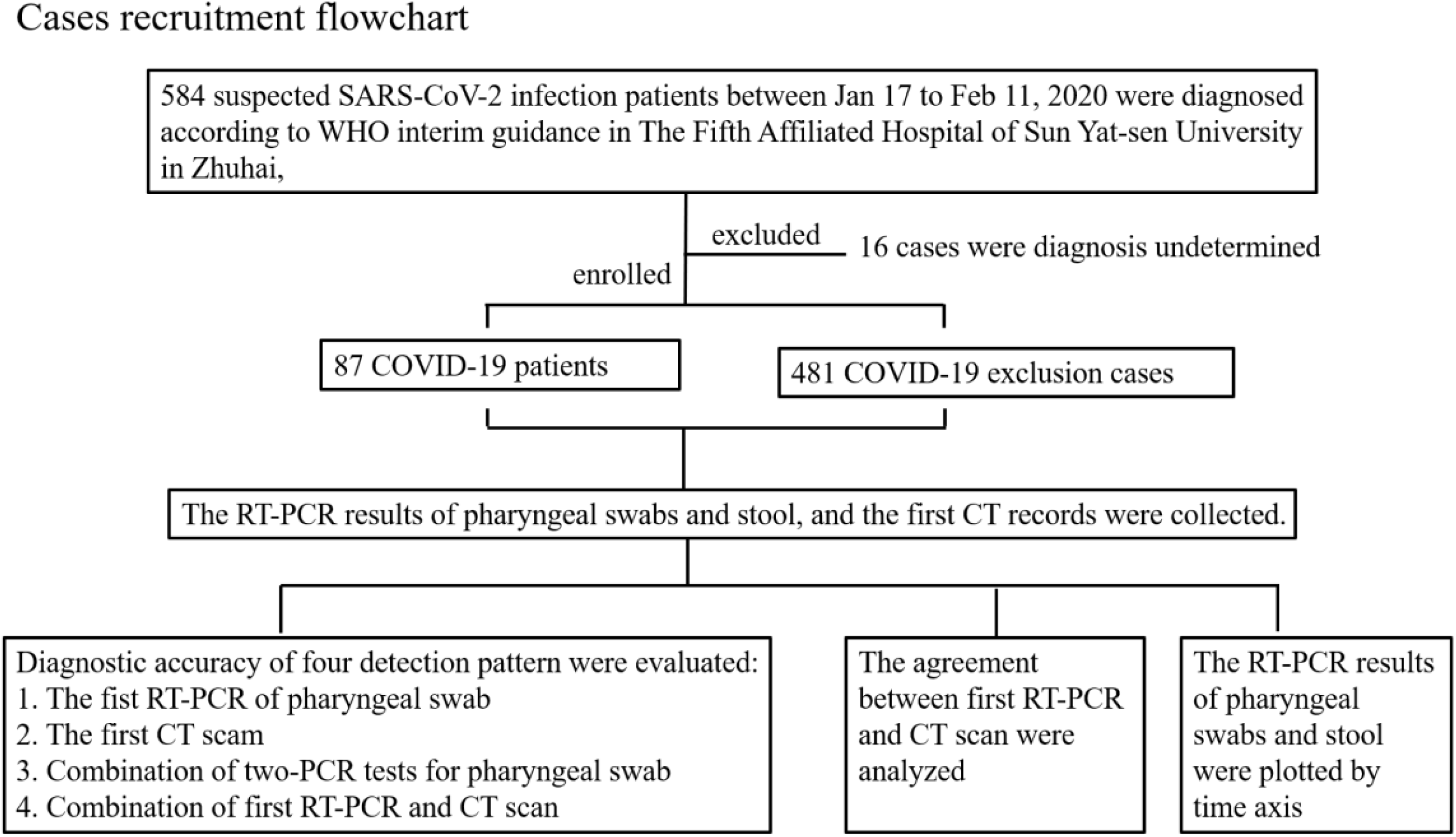
Flowchart for patient inclusion

**Figure 2.**
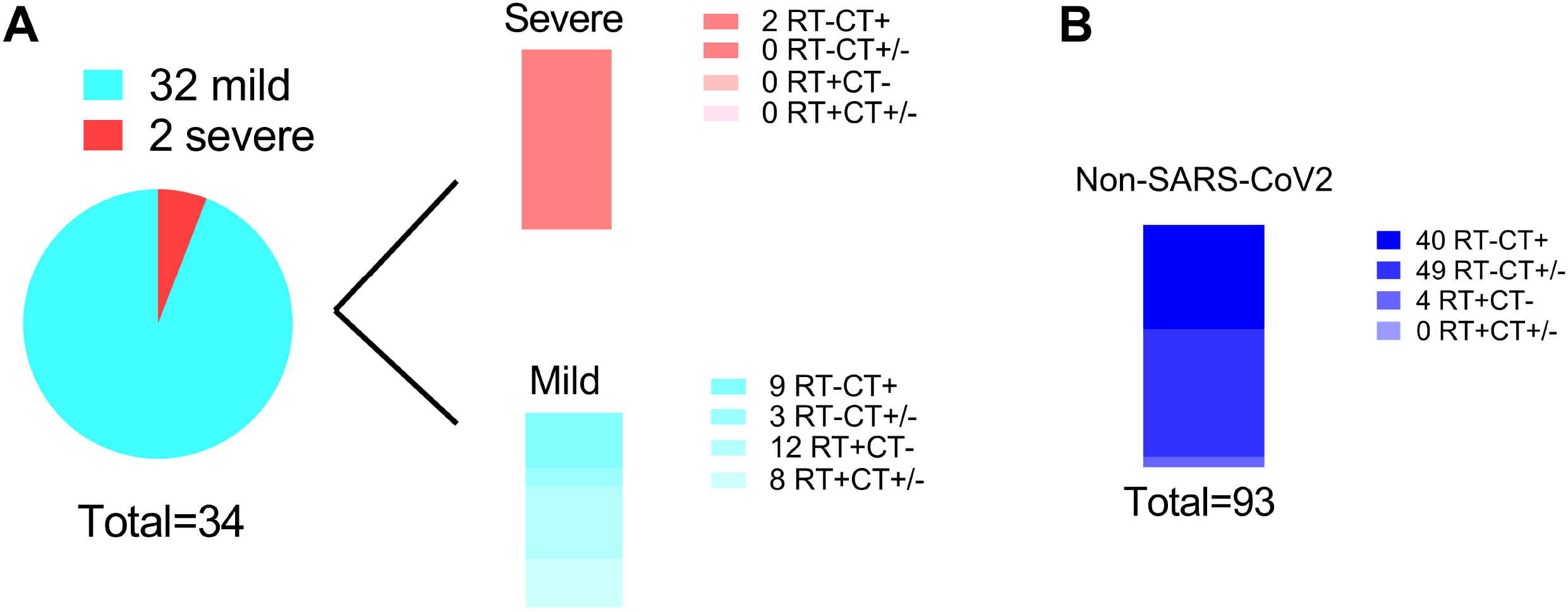
The analysis of 127 cases with disagreement of the first RT-PCR detection and CT scan. A. In COVID-19 patients. B. In non-COVID- 19 patients. In the legend, figures indicates the number of cases.

**Figure 3.**
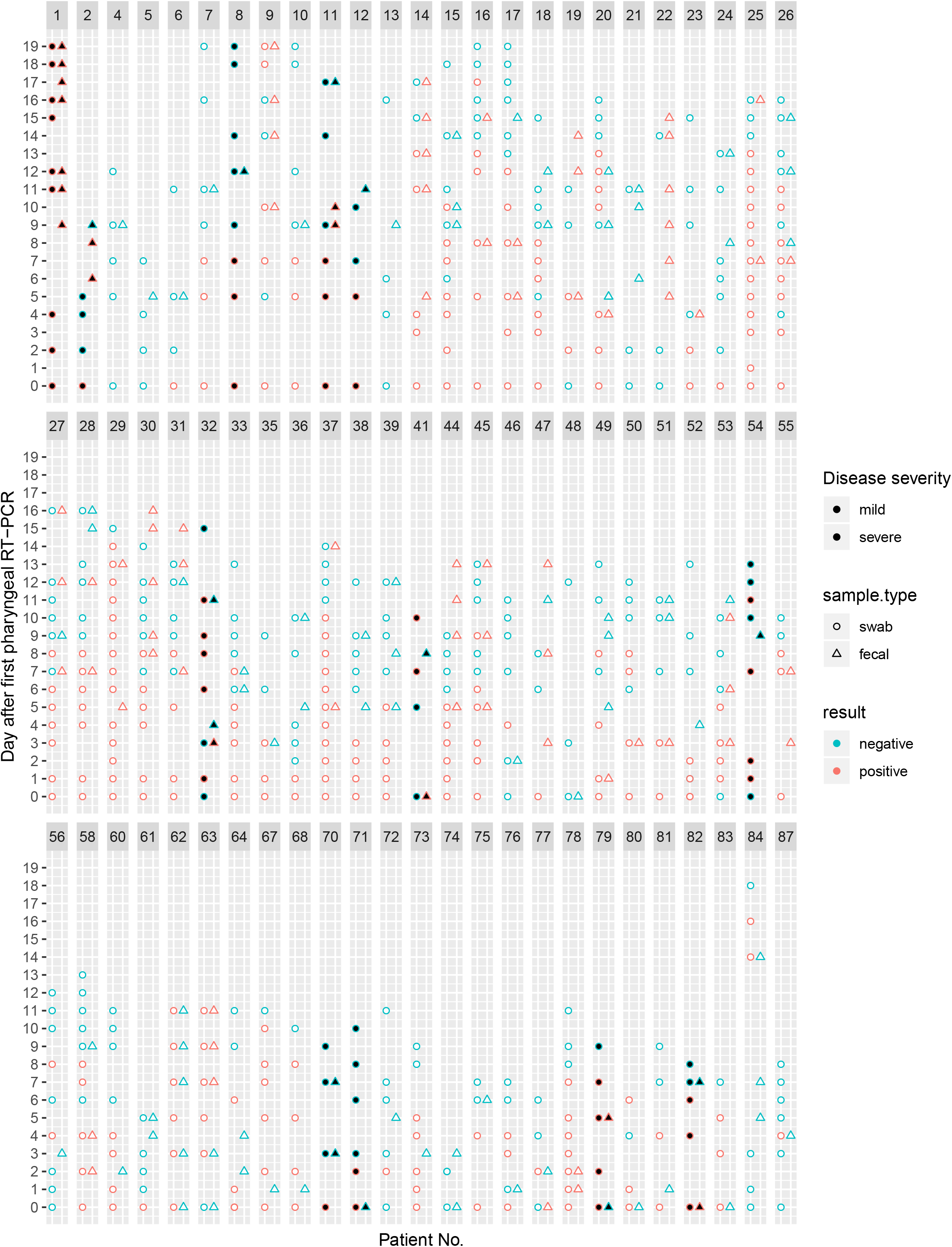
The qRT-PCR results were plotted along time axis in COVID-19 patients (n=75). Shapes and colors were used to represent sample types and results, respectively. Circle indicates pararenal swab and triangle indicates stool. Indigo indicates negative result and pale pink positive result. The dots filled with black represents severe patients. To the deadline of the study, 14 patients (patients No.2,3,4,5,10,11,12,17,23,32,33,54,62,63) had discharged from hospital.

The study was approved by The Fifth Affiliated Hospital of Sun Yat-sen University Ethics Committee and written informed consent was obtained from patients involved before enrolment when data were collected retrospectively.

### RNA Extraction and RT-PCR

The SARS-Cov-2 laboratory test assays were based on the previous WHO recommendation. The upper respiratory tract specimens (pharyngeal and nasal swabs) and stool were obtained from all the cases. Ensure each specimen collected has the name, gender and age of the patient as well as a serial number; any abnormality in the specimen should be noted.

RNA was extracted and tested by real-time RT-PCR with SARS-Cov-2 specific primers and probes according to instruction of Kit. The real-time RT-PCR was carried out in biosafety level 2 facilities at the clinic laboratory. If two targets (RdRp+, E or N +) tested positive by specific RT-PCR, the patients would be considered to be laboratory-confirmed.

Negative: no Ct value or Ct □ 40.

Positive: a Ct value < 37.

A Ct value between 37-40 is indeterminate. It is required confirmation by repeating. If, when repeated, the Ct value is < 37 the sample is positive, otherwise, it is negative.

### Chest CT

On admission, the chest CT images were detected among 365 patients. Of the 365 patients, typical and atypical chest CT findings were recorded. According to the Diagnosis and Treatment Program of 2019 New Coronavirus Pneumonia (trial sixth version), The typical findings of chest CT images were bilateral multiple lobular and subsegmental areas of consolidation, bilateral ground glass opacity and subsegmental areas of consolidation. Later chest CT images showed bilateral ground-glass opacity.

The findings with image features mentioned in Diagnosis and Treatment Program and interpreted by two radiologists are positive. No imaging abnormalities or exclusion of virus infection are defined as negative. That the abnormal imaging doesn’t accordance with the features in trial sixth version, but cannot rule out virological infection were defined as uncertain.

### Statistical analysis

Retrieved data were recorded into Microsoft ® Excel and analyzed. Continuous variables were expressed as median and range deviation. The independent sample t-test and one-sample t-test were utilized to compare significant differences among continuous data. Statistical analysis of agreement was performed using McNemar Chi-squared test. It was regarded as statistical significance when the value was less than 0.05. We used R (version 3.5.0) for all analyses.

## 3. Results

### 3.1 The common characteristics of the patients and specimen in this study

The patients involved in this study summed 568, including 87 COVID-19 and 481 non-COVID19. All of them were the local residents of Wuhan, had a recent travel to Wuhan or contacted with people from Wuhan. And they were all eligible for the epidemiology criteria of suspected cases, and received further medical isolation, examination and diagnosis. Among the patients, 58.2% were males. Throughout the whole spectrum of age, 15-59 years group generates 60.6% of all populations in this study, and 5.7% of patients were aged below 15 years (Table 1). In the disease severity, 83.9% (73 of 87 patients) were mild and 16.1% (14 of 87 patients)were severe.

**Table 1.**
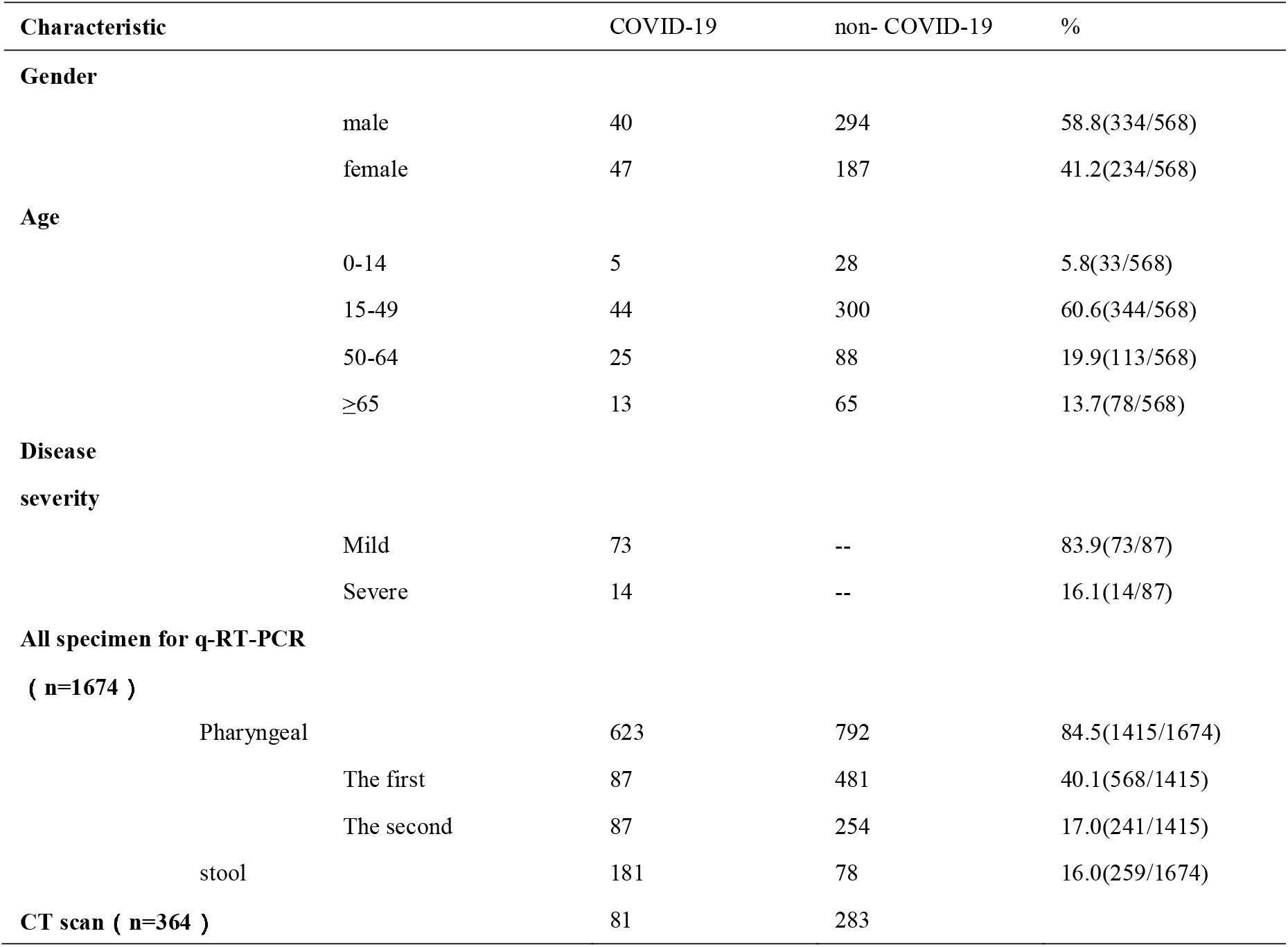
The common characteristics of the patients and specimen involved in this study

| Characteristic |  | COVID-19 | non- COVID-19 | % |
| --- | --- | --- | --- | --- |
| **Gender** |  |  |  |  |
|  | male | 40 | 294 | 58.8(334/568) |
|  | female | 47 | 187 | 41.2(234/568) |
| **Age** |  |  |  |  |
|  | 0-14 | 5 | 28 | 5.8(33/568) |
|  | 15-49 | 44 | 300 | 60.6(344/568) |
|  | 50-64 | 25 | 88 | 19.9(113/568) |
|  | ≥65 | 13 | 65 | 13.7(78/568) |
| **Disease severity** |  |  |  |  |
|  | Mild | 73 | -- | 83.9(73/87) |
|  | Severe | 14 | -- | 16.1(14/87) |
| **All specimen for q-RT-PCR** |  |  |  |  |
| **( n=1674 )** |  |  |  |  |
|  | Pharyngeal | 623 | 792 | 84.5(1415/1674) |
|  | The first | 87 | 481 | 40.1(568/1415) |
|  | The second | 87 | 254 | 17.0(241/1415) |
|  | stool | 181 | 78 | 16.0(259/1674) |
| **CT scan ( n=364 )** |  | 81 | 283 |  |

In the early stage of COVID outbreak, RT-PCR and CT scan were the main measure for screening SARS-CoV-2 infection. Hence we collected the results of the two methods in the 568patints in further. In the period of Jan 17^th^ to Feb 11^th^, total 1674 specimen from the 568 patients were detected by RT-PCR, which included 1415 pharyngeal and 259 stool (Table1). All patients after being admission received at least one RT-PCR detection for pharyngeal swabs, and 341 patients were subjected to the second RTPCR test for pharyngeal swabs. The stool detected by RT-PCR accounted to 259, 81 of which were from COVID-19 patients. Among the 568 patients, 364 patients were subjected to CT scan, 81 of which were COVID-19 patients. We looked through the case data retrospectively and found that 5 patients without CT scan were admitted to hospital complaining of epidemiology contact. Although they were eventually diagnosed, they had only mild symptoms of upper respiratory tract infection on admission and in the course of infection, and no CT scan had been performed.

### 3.2 The performance evaluation of the RT-PCR, CT scan and combination patterns in diagnosis of COVID-19

In the beginning of the SARS-CoV-2 outbreak, RT-PCR for pharyngeal and CT scan is the main screening strategy. Aiming to evaluate the property of the two methods in diagnosing SARS-CoV-2 infection, the performance indexes of RTPCR detection and CT scan were computed respectively. The results were shown in Table2. The sensitivity and specificity of RT-PCR for pharyngeal were 78.2% and 98.8%. positive predictive value and negative predictive value were 91.9% and 96.2. and Youden’s index was 0.770, which indicated overall performance. In the methodology evaluation of CT scan, criteria was defined firstly as an image diagnostic method. According the standard above, the data about the CT scan performance were evaluated. The sensitivity and specificity were 66.7% and 68.2%, lower than RT-PCR of pharyngeal. positive predictive and negative predictive value were 56.8% and 92.3% and Youden’s index was 0.343 (Table2).

Besides that, we also analyze the agreement of the two methods (The cases with uncertain CT results were not involved in statistical analysis due to statistical limitations). As is shown in Table3, there was statistically significance between the two methods (p<0.001). Moreover, there was statistically differences in the diagnosis of non-COVID-19 patients(p<0.001), but not in COVID-19 patients (p=0.734). The agreement was good to fair agreement (kappa value, 0.430), and the adjusted agreement was 72.8% in all patients.

Given that the time on receiving detection can cause influences on sensitivity, the time differences between RT-PCR for pharyngeal and CT scan were compared. The average time of nucleic acid screening was earlier than that of CT scan statistically (-0.7390days, -3625∼1.674) (Table 4). The COVID-19 group received an of CT scan later than the non-COVID-19 group in average (-1.7160,-5.185∼1.753 v.s. -0.4594, -2.391∼1.472, p=0.002) (Table 4).

**Table 2.** Performance of the RT-PCR test and CT scan in diagnosing SARS-CoV2 infection

|  |  |  |  |  |  |  |  |  |
| --- | --- | --- | --- | --- | --- | --- | --- | --- |
| Detection | Total | Sensitivity% | Specificity% | Positive<br>predictive<br>value% | Negative<br>predictive<br>value% | Positive<br>likelihood<br>ratio | Negative<br>likelihood<br>ratio | Youden's<br>Index |
| RT-PCR | 568 | 78.2(68/87) | 98.8(475/481) | 91.9(68/74) | 96.2(475/494) | 65.17 | 0.221 | 0.770 |
| CT<sup>a</sup> | 364 | 66.7(54/81) | 68.2(193/283) | 56.8(54/95) | 92.3(193/209) | 2.10 | 0.488 | 0.343 |
| RT&RT | 241 | 86.2(75/87) | 93.5(144/154) | 88.2(75/85) | 92.3(144/156) | 13.26 | 0.148 | 0.797 |
| RT&CT<sup>a</sup> | 370 | 91.4(74/81) | 66.8(189/283) | 62.2(74/119) | 97.9(189/193) | 2.75 | 0.129 | 0.581 |

<sup>a</sup> 60 cases manifested uncertain CT scan results, including 11 COVID-19 patients and 49 non-COVID-19 ones.

**Table 3.**
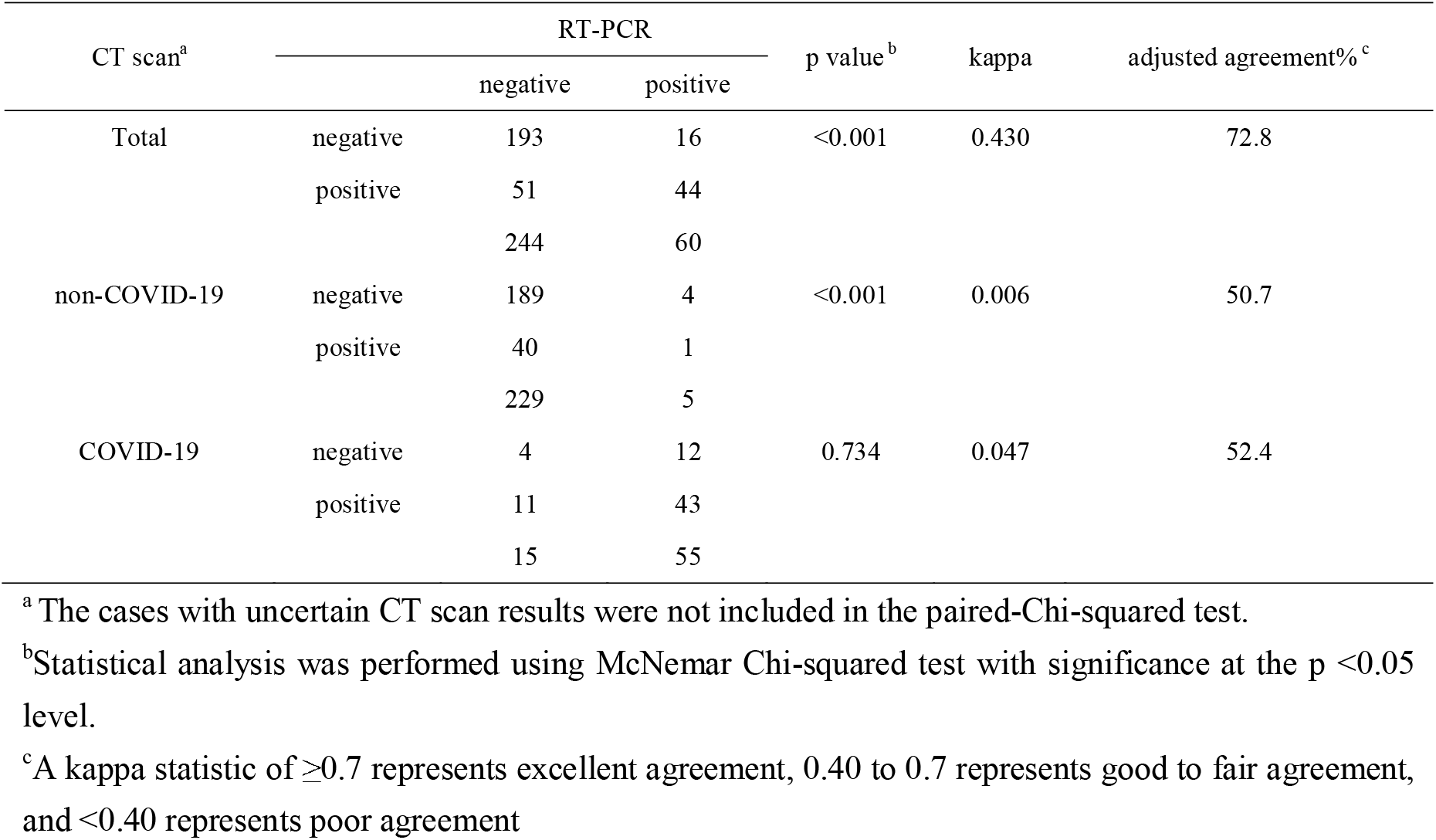
Comparison of RT-PCR and CT scan in diagnosis of SARS-CoV2 infection

**Table 4.** Statistics of the day differences between the first RT-PCR detection and CT scan

|  | Mean (95%CI) | p value* | p value# |
| --- | --- | --- | --- |
| All patients | -0.7390 (1.674~ -3.652) | <0.001 |  |
| Non-COVID-19 | -0.4594 (1.472~ -2.391) | <0.001 | 0.002 |
| COVID-19 | -1.7160 (1.753~ -5.185) | <0.001 |  |

The day differences between the first RT-PCR detection and CT scan were performed statistical analysis.
\*One-sample t test were used to test statistical significance between the mean of group and zero at the $p < 0.05$ level.

In purpose to explore the characteristics of the RT-PCR and CT scan in diagnosis of SARS-CoV-2 infections, 127 cases with inconsistent diagnosis by two methods were selected for further analysis, which is consisted of 34 COVID-19 patients and 93 non-COVID-19 cases (Fig.2). Notably mild cases count to 94.1% in COVID-19 suffers (32/34) (Fig2.A). 20 of the 32 cases showed RT-PCR positive & CT negative or RT-PCR positive & CT uncertain, suggested the priority of RT-PCR in identifying mild infections. There are still 12 cases with RT-PCR negative & CT positive or RT-PCR negative & CT uncertain, we are inclined to consider unqualified sampling or low viral load in early stage were responsible for the false discovery of RT-PCR. Only two inconsistent cases were severe infections (Fig2.A)., indicating both of the methods reached the good accordance. Besides that, we also focused the results pattern in 93 non-COVID 19, that mainly presented RT-PCR negative & CT positive (n=40) and RT-PCR negative & CT uncertain (n=49) (Fig2.B). That is 95.3% (89/93) patients possessed negative RT-PCR but abnormal CT results. We concluded that CT scan is an morphology detection, not pathogen identification, hence, it was difficult to differentiate SARS-CoV-2 from other viruses or pathogens accurately.

It is notable that both of the methods acquired the sensitivity less than 80% in screening SARS-CoV-22 infections, which is not ideal enough for the diagnosis of infectious diseases with severe consequences. To develop more appropriate detection scheme, the performances of combination RT-PCR and CT were evaluated in future. The 568 cases being subjected to first RT-PCR and second RT-PCR for pharyngeal and 341 being subjected to CT scan were analyzed. The performance indexes were shown in Table1. The results showed that the sensitivity of RT-PCR in parallel with CT scan was the highest(91.9%), which was higher than that of parallel with second nucleic acid (86.2%) (Table1). But the specificity of two nucleic acid detections was significantly higher than that of combination of nucleic acid and CT, suggesting that nucleic acid in parallel with CT was more appropriate to screen SARS-CoV-22 infections, and two nucleic acid tests for exclusion diagnosis maybe more suitable.

### 3.3 The value of nucleotide detection in stool was evaluated in COVID-19 patients

It is reported that alive virus can survive in stool of COVID-19 patients. According our data, 8.6% patients (Table1) cannot be identified by combination of RT-PCR for pharyngeal swab and CT scan. Since that, can the RT-PCR test for stool be an efficiency assisting examination? In this study, there were 75 COVID-19 cases subjecting to stool nucleotide detection. Therefore, all of the RT-PCR results from 75 case with stool nucleotide detection were plotted along time axis (Figure3). It was demonstrated that 35 cases had at least one positive results and the discovery rate is 46.7%. The stool presented earlier positive than the throat swab in two cases (patient No. 21, 38) (Figure3). Moreover, pharyngeal swab of No.21 patient had been always negative until the end of the study. There were 16 patients with remaining positive results of stool after two consecutive negative results of pharyngeal swabs (patient No.2,8,13,15,18,21,24,26,27,29,30,35,39,40,42,48) during their hospitalization (Figure3). Importantly, of the 14 discharged patients, two cases had stools being negative later than pharyngeal swabs (Patient No.2,10) (Figure3). The data above suggested that the detection of fecal nucleic acid could be employed to improve the discovery rate and might be developed as an indicator of monitoring and de-isolation.

## Discussion

SARS-CoV-2, a novel betacoronavirus are a major cause of symptomatic respiratory tract infection in all age groups worldwide^11,16,25^. Timely and accurate diagnosis of the virus enables appropriate treatment of infections. RT-PCR is widely deployed in diagnostic virology. In the case of a public health emergency, proficient diagnostic laboratories can rely on this robust technology to establish new diagnostic tests within their routine services before pre-formulated assays become available.

In our study, the sensitivity of RT-PCR was greater than that other reports (78% vs 30%-50%) in the first assay. The reasons for the high sensitivity of our series may include^26,27^: The first was samples, in some hospital, the patients’ nasopharyngeal or oropharyngeal swabs were collected for testing the SARS-CoV-2 separately. In our study, the detection specimen was the patient’s pharyngeal and nasal swabs combine, compared to the pharyngeal or nasal swab only, indicating higher sensitivity at initial screening. We show that the strategy for the detection of viral RNA in pharyngeal and nasal swabs used for SARS-CoV-2 diagnosis is not perfect. We also found that the virus are present in several stool swabs of patients when pharyngeal and nasal swabs detection negative. Based on the infected patients can potentially shed this pathogen through respiratory and fecal-oral routes, we applied test for oral and stool swabs which could greatly improved detection positive rate.

Among the 87 laboratory-confirmed cases, there were still 19 cases with RT-PCR results testing negative in the first assay. In the false negatives can be caused by poor sample quality, such as respiratory tract samples collected from the oropharynx; collection that is too early or late in the progression of the disease, in the early stages, the number of viruses in the body is not enough to be detected. samples that have not been properly stored, transported, or processed, SARS-CoV-2 coronavirus is RNA virus, which is prone to death and degradation. In the process of collecting samples and transporting them to the laboratory for testing, it takes a long time and the nucleic acid is easy to degrade, so it is not easy to detect positive. At last, technical factors, including virus mutation and PCR inhibition.

In our study, the sensitivity of RT-PCR was higher than that of the chest CT (78.5% vs 66.7%, respectively). The reasons for the relatively lower efficiency of chest CT is may include, 1) The major of COVID-19 patients was mild, in the early detection, there is no features or typical features of chest CT in our study. 2) We divided three groups in the 365 patients, one group is COVID-19 positive, the other group is COVID-19 negative. Our results support RT-PCR combine chest CT for first screening for COVID-19 for patients with clinical and epidemiologic features.

To date, two team from China have reported to succeed isolating alive SARS-CoV-2 in COVID-19 patients feces (data unpublished). According to these study, some of the provisions have been supplemented about strictly handling the patient’s secretions. Many coronaviruses can be transmitted through oral-fecal route by infecting intestines. SARS-CoV-2 belongs to lineage B, betacoronavirus (β-CoV) genus. The other member in this lineage is SARS-CoV, responsible for SARS outbreak in 2003 in China, can be detected in the stool of patients^28^. MERS, lineage C β-CoVs, has been proved existing in the feces^29,30^. For it is not too surprise to detect SARS-CoV-2 in stool. It is reported that SARS-CoV-2 RNA has been detected in the stool of a patient in the USA^16,17^. However, our study give an important hint that in a big portion of COVID-19 patients’ stool are there the RNA of SARS-CoV-2 and up to 16 patients presented positive stool after two negative pharyngeal swabs, which indicates stool nucleotide has potential role in monitoring infection as an supplement item.

Moreover, what deserve attention more is patients No.22, who had none positive pharyngeal swabs in all the stage of infection, but positive stool. Until the end of the observation, the patient were still under treatment. Although it can’t be absolutely excluded that pharyngeal presented false negative or shifting positive later, at least stool detection need be involved in examination for the strongly suspected persons with negative pharyngeal swab. Besides that, that stool remain positives in a later stage of infection suggests that fecal nucleic acid negative conversion should be included in the discharge criteria. In our center, there was no recurrence infection in 17 discharged patients, for the stool nucleotide had been detected before being out of hospitalization. Of course, we just provide the evidence the RNA in the stool. whether SARS-CoV-2 can be transmitted by oral to fecal need to be studied in future.

Combination of pharyngeal RT-PCR and chest CT with higher sensitivity is an reasonable option to screen SARS-CoV-2 infection patients. Two pharyngeal RT-PCR detections with higher specificity can be used in exclude diagnosis. RT-PCR has the more advantage in screening mild infection comparing to chest CT. RT-PCR of stool should be adopted to improve discovery rate and counted as an item for discharging from hospital. Our study shed light for the optional scheme of the clinical diagnosis and monitoring of SARS-CoV-2 infection.

## Data Availability

We declare that all data referred to in the manuscript and note links are available.

## Declaration of interests

All authors declare no competing interests

## Informed consent

None

## Notes

### Competing Interest Statement

The authors have declared no competing interest.

